# A Pilot Randomized Controlled Trial to Evaluate the Preliminary Efficacy of PAL-CHW-PDAC, a Digitally Enhanced CHW-led Intervention to Facilitate Stepped Palliative Care in Patients with Pancreatic Cancer

**DOI:** 10.64898/2026.05.20.26353748

**Authors:** Nikhil R. Thiruvengadam, Maud Celestin, Lizbeth Rivas, Arman Bahmani, Matthew Oroso, Nishita Matangi, Susanne Montgomery, Betty Ferrell

**Author notes:** **Correspondence to:** Nikhil Thiruvengadam, MD, 11234 Anderson Street, MC 1516, Loma Linda, CA 92354. **Author Contributions:** N.T., M.C, S.M., and B.F. designed the study; N.T. and L.R. enrolled patients in the study. N.T., A.B., and M.O. acquired the data; N.T., M.C., and S.M. interpreted the data and performed the statistical analysis. N.T. and B.F. wrote the manuscript. All authors discussed the data and the analysis methods and contributed to the manuscript.

## Abstract

**Background:** Pancreatic Ductal Adenocarcinoma will be the 2^nd^-most common cause of cancer mortality by 2030. It is associated with rapid deterioration, severe symptoms, and significant quality-of-life concerns. Using input from patients, family caregivers (FCGs), and provider stakeholders, we designed an intervention, PAL-CHW-PDAC, delivered by a community health worker that involves proactive symptom monitoring and management, care navigation, and disease education.

**Methods:** We conducted a pilot randomized controlled trial of 60 patients with newly diagnosed PDAC (within 2 weeks of diagnosis) and their caregivers at Loma Linda University Health from 09/2025 to 05/2026. Patients were randomized 1:1 to receive the PAL-CHW-PDAC intervention (6 CHW visits over 3 months) or an attention control. The control comparator involved receiving standard handouts and videos on pancreatic cancer, along with check-in visits with research staff. The primary outcome was symptom burden, defined using the NCCN/FACT Hepatobiliary Symptom Index. Secondary outcomes included quality of life (QoL) measured by the FACT-Hep and psychological distress (measured by the NCCN-Distress Thermometer). Caregiver outcomes included burden, preparedness, quality of life, and psychological distress.

**Results:** 60 out of 74 eligible (81%) were enrolled. The median age was 71, 60% of patients were Hispanic. 68% of patients presented with metastatic PDAC, 23% with borderline resectable disease and 9% with resectable PDAC. There was a trend towards improved symptom burden at 12 weeks (mean increase of 5.3 points vs. decrease of 3.2 points; p=0.093) with the intervention compared to the attention control. The intervention group also had improved psychological distress at 12 weeks (3.31 vs. 5.95, p=0.01), caregiver psychological distress (3.26 vs. 6.86, p<0.001) and caregiver preparedness (2.92 vs. 2.11) at 12 weeks. Telehealth utilization for symptom-focused visits improved with the intervention (82%) compared to the control. (14%, p=0.01) Hospice utilization also improved with the intervention (41% vs 7%, p-0.12).

**Conclusions:** A pilot RCT of the PAL-CHW-PDAC intervention demonstrated preliminary efficacy with a trend towards improved symptom burden, psychological distress, and caregiver psychological distress and preparedness. A larger definitive clinical trial is needed to understand the impact of this promising intervention. ClinicalTrials.gov number, NCT07591571

## Background

PDAC has become the 3rd most common cause of cancer mortality, projected to become the 2nd largest contributor to cancer mortality by 2030.^1^ The overall 5-year survival is 12%, with poor survival (44%) even in patients with local-stage disease.^2^ Patients with PDAC typically present with late-stage disease^3^ resulting in high symptom burden, poor quality of life (QOL), and psychological distress from awareness of a poor prognosis. Multiple clinical trials have demonstrated that Early Palliative Care (EPC), when initiated within 2-4 weeks of diagnosis, can improve physical, social, emotional, and functional QOL in PDAC.^4, 5^ It also increases advanced care planning (ACP) and reduces overall healthcare costs.^6^ Early PC (EPC) can reduce the morbidity and symptom burden associated with PDAC.

However, only 7.7% of patients with locally advanced PDAC received supportive and palliative care (PC), mostly occurring near the end of life. ^7^ Late referral to PC is not ideal as PC benefits cannot be fully realized, leading to unnecessary symptom burden, morbidity, and higher healthcare costs.^8, 9^ In contrast, PC encounters within 30 days of diagnosis reduced ED visits, the rate of ICU admission, and overall healthcare costs.^6^

Novel modes of delivering PC, including stepped PC models, have been developed in which visits are “stepped up” as the disease or symptoms worsen. A recent RCT demonstrated that stepped PC was non-inferior to early scheduled PC in advanced lung cancer, with regard to QOL, required fewer PC visits, and was more scalable.^10^ Furthermore, a multi-center RCT demonstrated that telehealth-based EPC was non-inferior to in-person care for delivering EPC in lung cancer.^11^ Despite this, in our cohort of 244 patients with PDAC, less than 6% used telehealth for PC or symptom-focused oncology visits.^12^ We found that low digital literacy, older age (median age of 73), language barriers, and rural residence were all associated with lower odds of using telehealth.

We thus developed We developed a novel CHW-led intervention (PAL-CHW-PDAC) in 2024 focused on symptom burden. The intervention facilitates stepped PC, including symptom management, care navigation, and patient education paired with an SMS-based digital application to assess patients’ symptom burden weekly by administering patient-reported outcomes (PROs). Our intervention was feasible and acceptable to patients, resulting in improvements in symptom burden (measured by the NCCN/FACT Hepatobiliary Symptom Index, (NFHSI-18) as our primary outcome at 12 weeks. ^13^ Other improved secondary outcomes included psychological distress (measured by NCCN Distress Thermometer) and advance directive completion.

In this study, we conducted a pilot randomized controlled trial to assess the preliminary efficacy of the PAL-CHW-PDAC intervention in PDAC patient-caregiver dyads.

## Methods

This RCT was performed in a single center, Loma Linda University Medical Center (LLUMC), in Loma Linda, CA. (ClinicalTrials.gov NCT07591571). The study was reported according to the CONSORT guidelines for RCTs. This study was funded by the DOD/CDMRP. All authors had access to the study data and reviewed and approved the final manuscript. The local institutional review board approved the study (IRB# 5250314), as well as the Office for Human Research Oversight at the US Army Medical Research and Development Command (Protocol Number E05835.3a, as the funder of the study). Written and informed consent was obtained from all subjects before their inclusion in the study.

### Study Population

We included patients newly diagnosed with PDAC (within 2 weeks of diagnosis) and their FCGs. We enrolled patient-FCG dyads for this study. Additional eligibility criteria include: 1) Age 18 years or older, 2) Ability to read or understand English, or Spanish, 3) Access to a SMS-capable phone.

### PAL-CHW-PDAC Intervention

We previously published the design and structure of the PAL-CHW-PDAC intervention. In Table 1, we summarize the contents of the intervention, which includes 6 visits over 12 weeks.

**Table 1.**
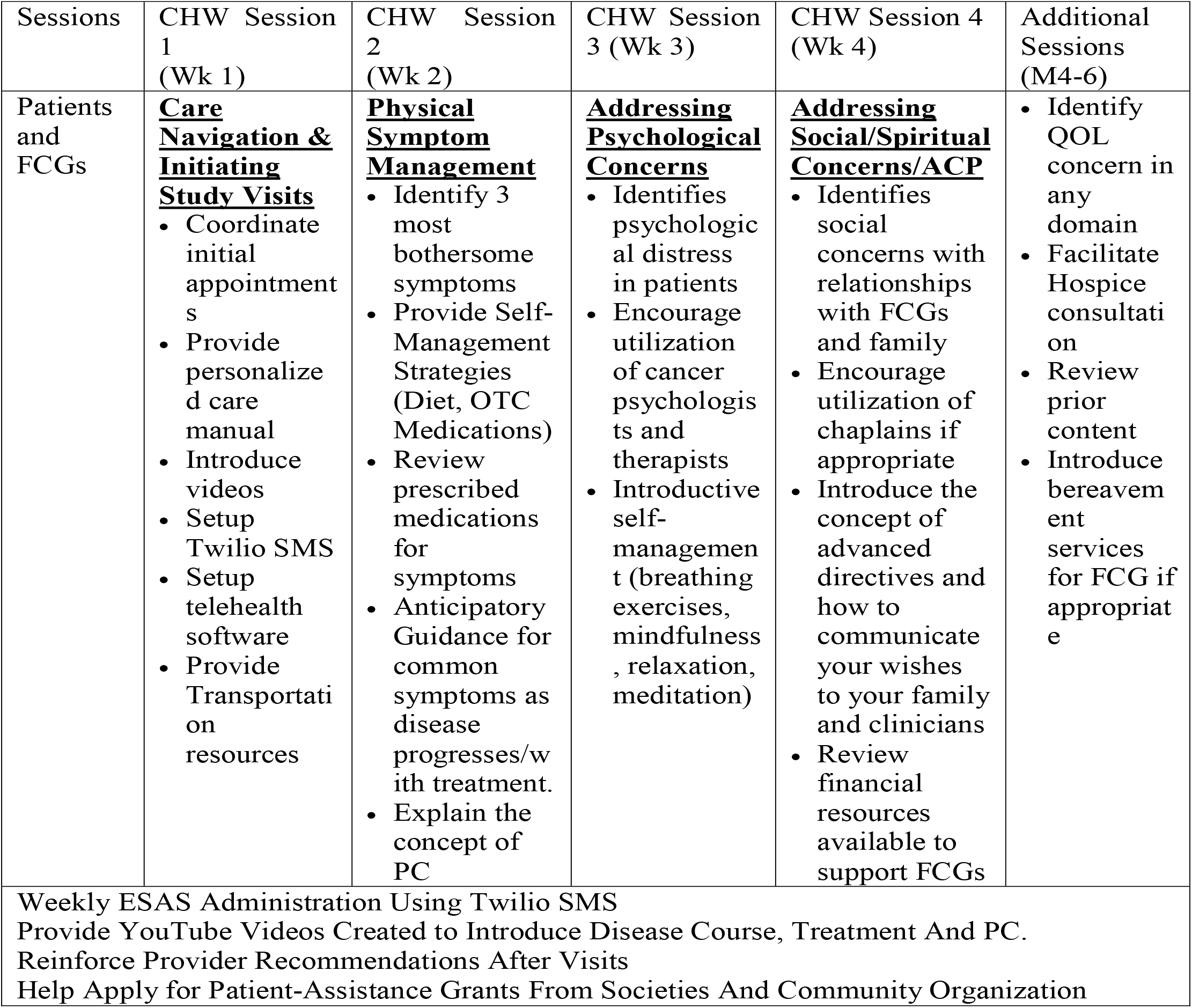
Summary of PAL-CHW-PDAC Intervention.

| Sessions | CHW Session 1 (Wk 1) | CHW Session 2 (Wk 2) | CHW Session 3 (Wk 3) | CHW Session 4 (Wk 4) | Additional Sessions (M4-6) |
| --- | --- | --- | --- | --- | --- |
| Patients and FCGs | **<u>Care Navigation & Initiating Study Visits</u>**<br>• Coordinate initial appointments<br>• Provide personalized care manual<br>• Introduce videos<br>• Setup Twilio SMS<br>• Setup telehealth software<br>• Provide Transportation resources | **<u>Physical Symptom Management</u>**<br>• Identify 3 most bothersome symptoms<br>• Provide Self-Management Strategies (Diet, OTC Medications)<br>• Review prescribed medications for symptoms<br>• Anticipatory Guidance for common symptoms as disease progresses/with treatment.<br>• Explain the concept of PC | **<u>Addressing Psychological Concerns</u>**<br>• Identifies psychological distress in patients<br>• Encourage utilization of cancer psychologists and therapists<br>• Introductory self-management (breathing exercises, mindfulness, relaxation, meditation) | **<u>Addressing Social/Spiritual Concerns/ACP</u>**<br>• Identifies social concerns with relationships with FCGs and family<br>• Encourage utilization of chaplains if appropriate<br>• Introduce the concept of advanced directives and how to communicate your wishes to your family and clinicians<br>• Review financial resources available to support FCGs | • Identify QOL concern in any domain<br>• Facilitate Hospice consultation<br>• Review prior content<br>• Introduce bereavement services for FCG if appropriate |
| Weekly ESAS Administration Using Twilio SMS<br>Provide YouTube Videos Created to Introduce Disease Course, Treatment And PC.<br>Reinforce Provider Recommendations After Visits<br>Help Apply for Patient-Assistance Grants From Societies And Community Organization |  |  |  |  |  |

**Table 2.**
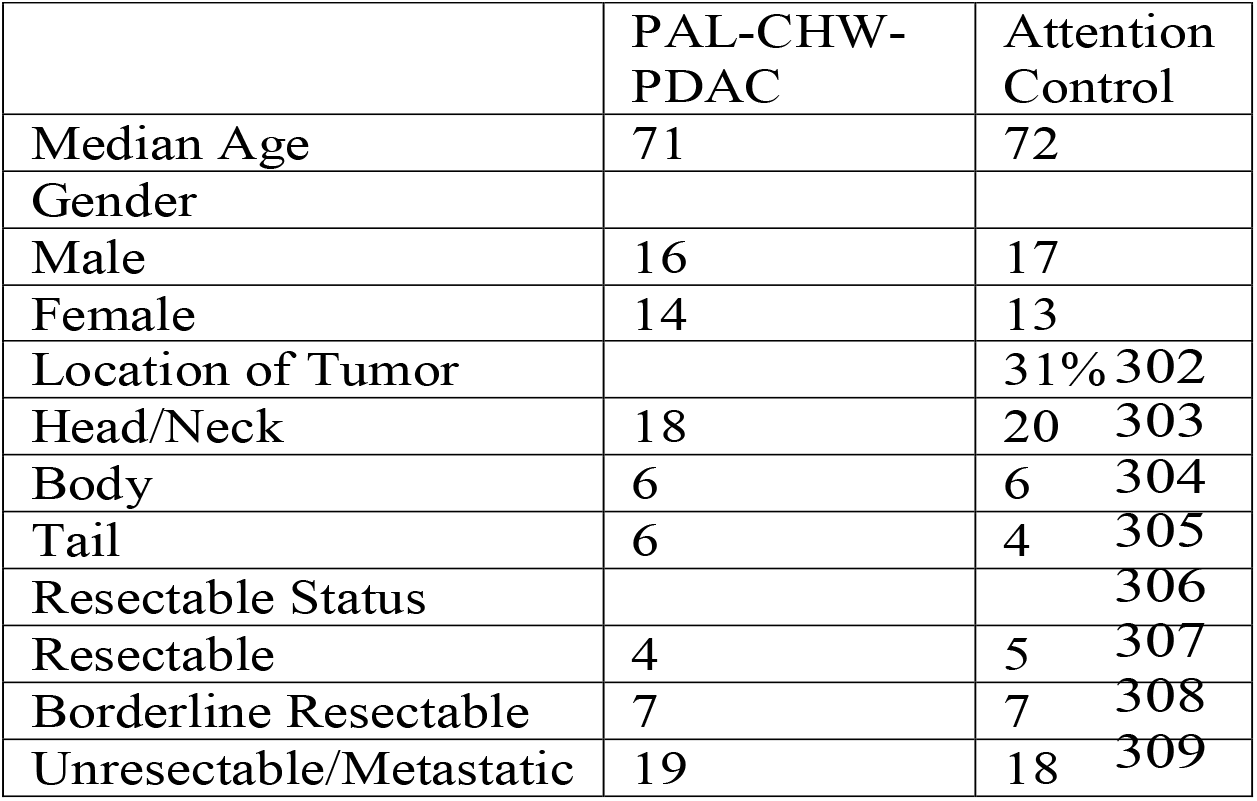
Baseline Characteristics of Both Groups.

**Table 3.** Resource Utilization at 3 months in Pilot RCT.

| Table 3. Resource Utilization at 3 months in Pilot RCT<sup>317</sup> |  |  |  |
| --- | --- | --- | --- |
|  | PAL-CHW-PDAC | Attention Control | p-value |
| Time to First Oncology Visit | 9±6 days | 15±8 days | p=0.16 |
| Time to Starting Treatment | 16±8 days | 21±6 days | p=0.20 |
| Telehealth Utilization for Symptom Focused Visits | 82% | 14% | p<0.001 |
| Complete Germline Genetic Testing | 91% | 44% | p<0.001 |
| ACP Completion | 86% | 31% | p<0.001 |
| Hospice Utilization | 41% | 7% | p=0.003 |

### Usual Care

Patients in both groups will receive ASCO guideline-based care at all 3 sites, which involves patients who receive a diagnosis of PDAC at the institution (with positive cytology from an endoscopic biopsy), referred to medical oncology, surgical oncology, and PC. PC referrals at the time of diagnosis are standard practice at all three sites.

### Attention Control

This study used an attention control design as the control comparator because it is considered one of the most rigorous in behavioral interventions.^14^ Importantly, an attention-control design accounts for the potential effects of receiving attention from a CHW. It also ensures that participants feel equally valued in each assigned group, minimizing study attrition and reducing the potential for inflation of the PAL-CHW-PDAC arm results. The study’s attention control condition included: 1) provision of the Pancreatic Cancer Action Network’s Education Packet^15^ (available freely as a PDF or written booklet); 2) being provided the link to digital videos from the National Pancreas Foundation about PDAC and 3) weekly telehealth (Doximity or Teams) encounters with the CRC to answer questions about the booklet in the first month, followed by monthly visits in months 2 through 3. The number of study visits will be the same in the attention control and interventional arms.

### Study Outcomes

The primary outcome was the change in symptom burden from baseline to 12 weeks using the NHFSI-18.^16^ Secondary patient outcomes included QoL (measured by the FACT-Hep^16^) and Psychological Distress (measured by the NCCN Distress Thermometer^17^). Caregiver outcomes include the change from baseline to 12 weeks in burden (measured by Montgomery-Borgatta Caregiver Burden scale^18^), preparedness (Preparedness for Caregiving Scale^19^), Psychological Distress (measured by the NCCN Distress Thermometer^17^), and QoL (measured by COH-QOL-Family Instrument^20^). Other secondary outcomes were healthcare utilization, specifically the utilization of telehealth, germline genetic testing and hospice.

### Sample Size

Prior statistical studies have found that sample sizes of 20 to 30 per arm in a pilot RCT are adequate to use to power definitive clinical trials of interventions.^21^ Thus we used a sample size of 30 per arm in our study.

### Statistical Analysis

Demographics and oncologic characteristics were connected. Instrument scores were examined for normality. Some outcomes were normally distributed and required transformation before performing a repeated-measures ANCOVA. Normalizing transformations will be considered if necessary to meet the assumptions of the statistical methods to be used. Participants in the treatment arm will be compared with the attention control participants on demographic and clinical characteristics, including site and disease stage (resectable/borderline resectable vs. metastatic), to provide a full description of each group and the overall sample.

To test the effect on patients’ symptom burden, a 2×2 repeated-measures ANCOVA was performed, with within-group variables being symptom burden scores measured by the NFHSI at 4 and 12 weeks. The between-group measures were comparisons between study arms (attention control vs intervention), with the baseline symptom burden score serving as a covariate.

All statistical analyses were performed using STATA 17 statistical package (StataCorp LP, College Station, TX). A p-value < 0.05 was considered statistically significant for all analyses.

## Results

Between September 1, 2025, and May 1, 2026, 71 patients were screened for inclusion in the study, and 60 were enrolled. 59 caregivers were enrolled as well. (1 patient lacked a family caregiver but was included in the study). 5 patients in the attention control arm and 4 in the intervention arm died before the 12-week endpoint and thus are not included in the analysis of that endpoint.

The median age was 71, 60% of patients were Hispanic. 68% of patients presented with metastatic PDAC, 23% with borderline resectable disease and 9% with resectable PDAC. There were no differences in baseline characteristics between intervention and attention control arms.

Regarding the primary outcome, there was a trend towards improved symptom burden at 12 weeks (mean increase of 5.3 points vs. decrease of 3.2 points; p=0.093) with the intervention compared to the attention control. (Figure 1) The intervention group also had reduced psychological distress at 12 weeks (3.31 vs. 5.95, p=0.01), reduced caregiver psychological distress (3.26 vs. 6.86, p<0.001), and increased caregiver preparedness (2.92 vs. 2.11, p=0.02). No difference was seen in FACT-Hep at 12 weeks between the intervention and attention control groups (96 vs 92) or in caregiver QoL at 12 weeks (mean 5.32±1.71 vs 5.22±1.55).

**Figure 1.**
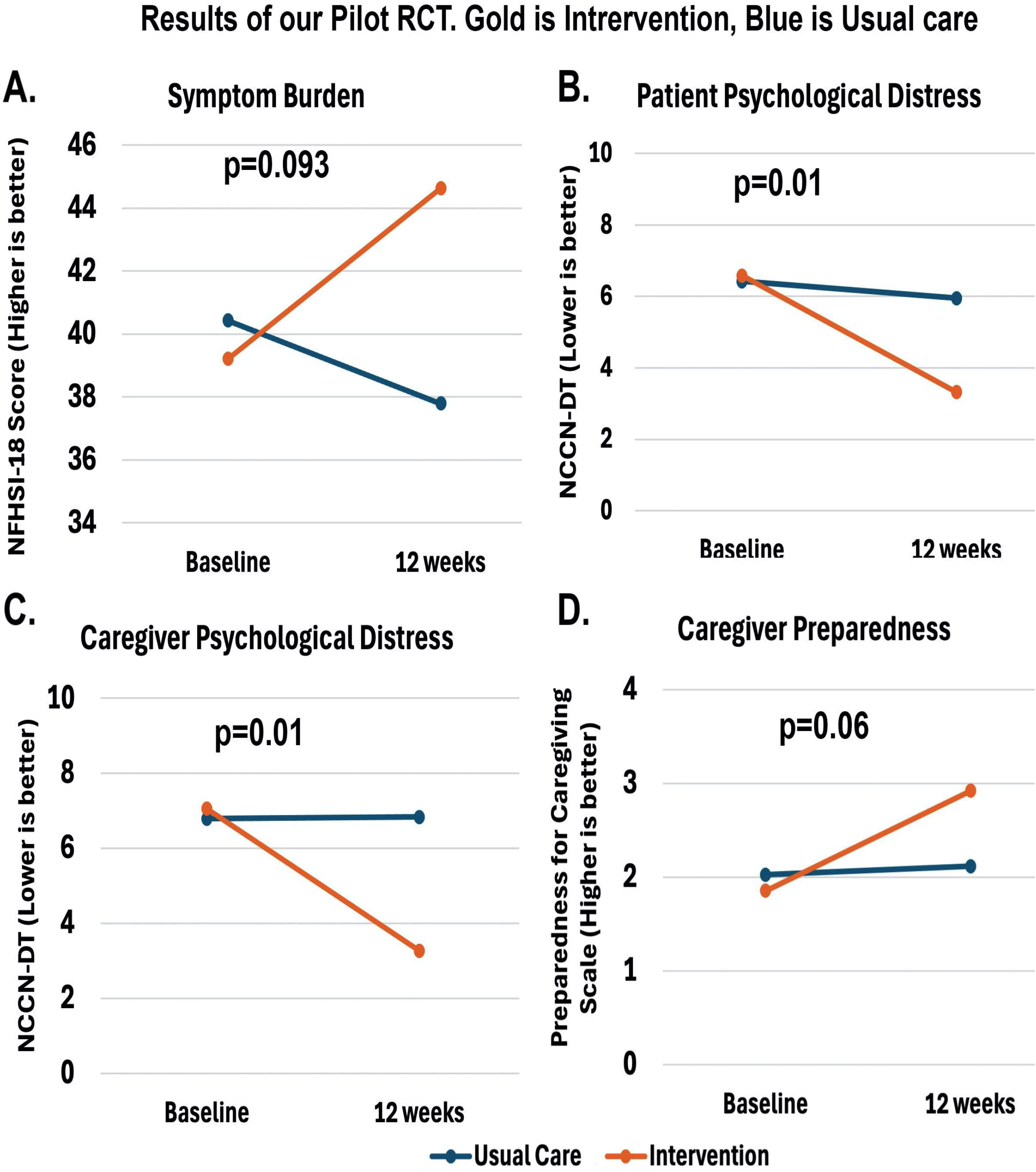
Comparison of the intervention vs attention control on patient symptom burden (A), patient psychological distress (B), caregiver psychological distress (C) and caregiver preparedness.

Regarding resource utilization, telehealth utilization for symptom-focused visits improved with the intervention (82%) compared to the control. (14%, p<0.001) Rates of completion germline genetic testing was significantly higher than in the intervention group (91%) compared to the attention control (44%, p<0.001). Similarly, rates of ACP completion were significantly higher in the intervention group than in the attention control group (intervention 86% vs. control 31%, p<0.001). Hospice utilization also improved with the intervention (41% vs 7%, p < 0.003).

Regarding delays in care, no statistically significant difference was observed in the time to first oncology visit or the time to start treatment. However, in terms of acute care utilization (emergency room visits/hospitalizations), the rate was lower at 12 weeks in the intervention arm (mean 2.4±1.4) compared to the attention control arm (mean 3.7±1.9; p=0.04).

## Discussion

In this study, we are the first to report the results of a pilot RCT comparing a novel CHW-led intervention, PAL-CHW-PDAC, to an attention control for patients with newly diagnosed PDAC and their family caregivers. We demonstrated a trend towards reduced symptom burden in this high-need population, with reductions in patient psychological distress and increased utilization of supportive services. Importantly, a significantly larger proportion of intervention patients utilized a telehealth-based symptom-focused visit, which is critical given the shortage of PC providers and the challenge in connecting older and rural patients with tertiary-center-based PC clinics. Furthermore, we demonstrated that the intervention improved caregiver outcomes, including both psychological distress and caregiver preparedness. Our results are promising and suggest that a larger definitive clinical trial is warranted to assess the effectiveness of the intervention and its feasibility for implementation in clinical practice.

Patients with PDAC experience a substantial symptom burden from the time of diagnosis. In our prior prospective cohort study of 111 patients with newly diagnosed PDAC, 36% reported increased symptom burden driven primarily by abdominal pain, fatigue, nausea/vomiting, and early satiety.^22^ More studies have demonstrated a rapid deterioration in symptom burden and quality of life after starting treatment. For patients receiving palliative chemotherapy, baseline QoL was significantly impaired, and 39% had deterioration in QoL by 3 months and 59% by 6 months.^23^ Indeed, symptoms related to PDAC have been classified into three main categories: pain-related symptoms, gastrointestinal symptoms, liver dysfunction-related symptoms (due to biliary obstruction).^24^ A prior study demonstrated that early specialty palliative care within 3 weeks of diagnosis with scheduled visits every 2 weeks reduced symptom burden and improved QoL in PDAC.^25^ For this reason, ASCO guidelines recommend early PC in PDAC and our institution started placing PC referrals at diagnosis. However, due to limited PC provider availability and patients prioritizing starting treatment and oncologic care, PDAC patients would see a specialty PC provider 67 days after diagnosis. The PAL-CHW-PDAC intervention was able to overcome these limitations by providing a focused education curriculum, so patients and caregivers understand the importance and purpose of PC. It demonstrated a signal towards reducing symptom burden through proactive weekly symptom monitoring, with escalation to PC visits for patients with new or untreated symptoms. The CHW also reinforces provider recommendations and providers’ self-management strategies. With a larger sample size, we can assess if these promising results reflect true improvement in symptom burden.

CHWs also represent the ideal workforce to engage and help facilitate early and stepped PC. While there is a severe shortage of PC providers, the number of CHWs working in the US is expected to grow by 13% annually, representing the fastest-growing workforce in healthcare.^26, 27^ CHWs are growing faster in California, with an annual increase of 25%, with almost 25,000 trained and available in 2025.^28^ This is largely due to improved reimbursement. In 2024, Medicare began reimbursing CHW services if they addressed social determinants of health.^29^ Regarding funding, more than 37 states have passed bills providing CHW service coverage through Medicaid and other managed care programs.^30^ In California, starting in 2022, Medi-Cal has covered and reimbursed CHW services, and a subsequent bill in 2025 streamlined their supervision and reimbursement.^31^ We demonstrated that with a structured training program, careful supervision and oversight and structured integration into clinical teams, CHWs can help bridge the gap in PC care.

Another unique aspect of our intervention is that the CHW helped patients and caregivers utilize technology particularly telehealth services. Telehealth has been an attractive modality to deliver PC as it eliminates bottlenecks such as available clinic space, the lack of PC providers close to many patients in the community and also allows for more rapid visits in response to symptoms. In lung cancer, a multi-center RCT demonstrated that telehealth-based EPC was non-inferior to in-person care for delivering early.^11^ However, no telehealth-based PC interventions exist for PDAC and we had previously demonstrated low rates of utilization of telehealth for PC in our center.^12^ We found low rates of digital literacy, older age (median age of 73), language barriers, and rural location were all associated with lower odds of using telehealth. We were able to overcome these barriers as CHWs were able to go to the patient’s home and help them utilize existing devices for completing symptom-focused telehealth visits.

Another unique aspect of our intervention is we also addressed the needs of family caregivers. Importantly, we provided focused education and connected them to resources including low-cost behavioral health resources in the community and support groups. We demonstrated that this education improved caregiver preparedness and also reduced rates of psychological distress.

While this study has many strengths it does have several weaknesses. The main weakness is the small sample size as it is indeed just a pilot RCT. Thus, we lacked the statistical power to assess many outcomes. However, we did demonstrate that randomization in this high-acuity population was feasible. Another limitation is the intervention was delivered in a single center with a small number of CHWs (2). It remains to be seen if this can be scaled to multiple centers and multiple CHWs, which highlights the importance of implementation studies and assessing intervention fidelity when it is scaled to other sites. The final limitation is that there could have been contamination since the same providers were caring for both the intervention and attention control groups. However, the intervention is working with the CHW and referrals to all providers are standardized between both groups.

In conclusion, our pilot RCT demonstrated that PAL-CHW-PDAC shows promise in improving patient and caregiver outcomes in PDAC. A larger definitive clinical trial is needed to study its impact along with implementation studies to see if it can be scaled to other communities and centers.

## Data Availability

No data produced in the present study will be available due to the sensitive nature of the data.

## Abbreviations used in this paper

CI: (Confidence Interval)
CHW: (Community Health Worker)
PC: (Palliative Care)
PDAC: (Pancreatic Ductal Adenocarcinoma)
RCT: (randomized control trial)
PROM: (Patient-Reported Outcome Measure)

